# An Application of Nowcasting Methods: Cases of Norovirus during the Winter 2023/2024 in England

**DOI:** 10.1101/2024.07.19.24310696

**Authors:** Jonathon Mellor, Maria L Tang, Emilie Finch, Rachel Christie, Oliver Polhill, Christopher E Overton, Ann Hoban, Amy Douglas, Sarah R Deeny, Thomas Ward

## Abstract

**Background:** Norovirus is a leading cause of acute gastroenteritis, adding to strain on healthcare systems. Diagnostic test reporting of norovirus is often delayed, resulting in incomplete data for real-time surveillance.

**Methods:** To nowcast the real-time case burden of norovirus a generalised additive model, semi-mechanistic Bayesian joint process and delay model, and Bayesian structural time series model including syndromic surveillance data were developed. These models were evaluated over weekly nowcasts using a probabilistic scoring framework.

**Results:** Modelling current cases clearly outperforms a simple heuristic approach. Models that harnessed a time delay correction had higher skill, overall, relative to forecasting techniques. However, forecasting approaches were found to be more reliable in the event of temporally changeable reporting patterns. The incorporation of norovirus syndromic surveillance data was not shown to improve model skill in this nowcasting task, which may be indicative poor correlation between the indicator and norovirus incidence.

**Interpretation:** Analysis of surveillance data enhanced by nowcasting reporting delays improves understanding over simple model assumptions, which is important for real-time decision making. The structure of the modelling approach needs to be informed by the patterns of the reporting delay and can have large impacts on operational performance and insights produced.

## Introduction

Norovirus is a gastrointestinal RNA virus causing symptoms of nausea, vomiting and diarrhoea. Norovirus often causes outbreaks in enclosed settings, such as hospitals, care homes and nurseries [1], causing a substantial burden on health systems, particularly over winter [2] [3]. Norovirus transmission was limited during lockdown periods of the SARS-CoV-2 pandemic response, followed by resurgent spreading when normative population mixing patterns resumed to pre-pandemic levels [4]. Norovirus is a constantly evolving pathogen with antigenic drift and shift [5] leading to strain replacement events periodically [6] [7] and resulting in short-lived immunity. These events cause large outbreaks and elevated transmission, highlighting the importance of monitoring and improving the timeliness of insights for taking public health action.

Norovirus surveillance in England uses data from multiple national surveillance systems. These include norovirus positive laboratory reports from confirmed cases, of which a subset undergo molecular typing, as well as notifications of outbreaks [8]. There is a reporting delay between diagnostic test administration and reporting to the national surveillance data, partially attributable to norovirus not being a Schedule 2 notifiable causative agent in legislation [9]. Due to this lag, the national official statistics surveillance reports truncate the time period shown by one week to remove partially complete data [8].

Norovirus is an excellent candidate for the application of nowcasting methods due to the inherent delay in case reporting as a non-priority pathogen. Research has been conducted on short term projections using statistical methods [10] [11], though there is limited exploration of correcting for time delays in norovirus cases. Norovirus incidence is highly stochastic, with a partially seasonal pattern and high heterogeneity between localised outbreaks and national trends, making it challenging to predict. Building on nowcasting research applied during the SARS-CoV-2 pandemic [12] [13] modelling can be explored to improve understanding of the real-time norovirus dynamics.

In this paper, we explored the reporting delay for norovirus cases in England over the 2023/2024 winter. We evaluated a range of methods for nowcasting using different model structures, guide signals, and assumptions about data completeness to consider the trade-offs between different approaches applicable to norovirus and beyond.

## Methods

### Data

#### Norovirus Cases

Individual test results were extracted from the Second Generation Surveillance Service (SGSS) database in UKHSA (UK Health Security Agency) [14] for positive norovirus tests conducted in England. The database only stores information on positive laboratory test results uploaded by frontline diagnostic laboratories, with a sampling bias towards health and social care settings. We deduplicated tests to keep the first test per patient infection episode. Under the legislation positive norovirus diagnostic tests are required to be notified to the UKHSA, but not required within 7 days of testing [15]. Cases followed a day-of-week periodicity shown in Supplementary Figure 1.

In this analysis we focused on two main time events for each test. Firstly, the specimen date *t* defines when the specimen was collected from the infected individual for testing. Secondly, the report date *t*_*r*_ defines the date the record is ingested into SGSS, thereby notifying UKHSA of the norovirus case through national surveillance. As symptom onset dates are not reported, the specimen date is the most epidemiologically relevant event. Despite being impacted by time to treatment, the specimen date gave the least delayed representation of the epidemic’s progression compared with other available time events for each test. The difference between report date and specimen date *d= t*_*r*_ −*t* is thereporting delay.

To model the epidemic and corresponding delay distributions, we aggregated the data by *t* and *d* to construct a so-called data “reporting triangle” [16], illustrated in Figure 1. The reporting triangle is an array with elements, for and *n*_*t,d*_, for *t* ∈[1, *T*] and *d* ∈ [0, *D*], where *T* is the maximum length of the specimen date time series *t* and *D* is the maximum reporting delay.

**Figure 1.**
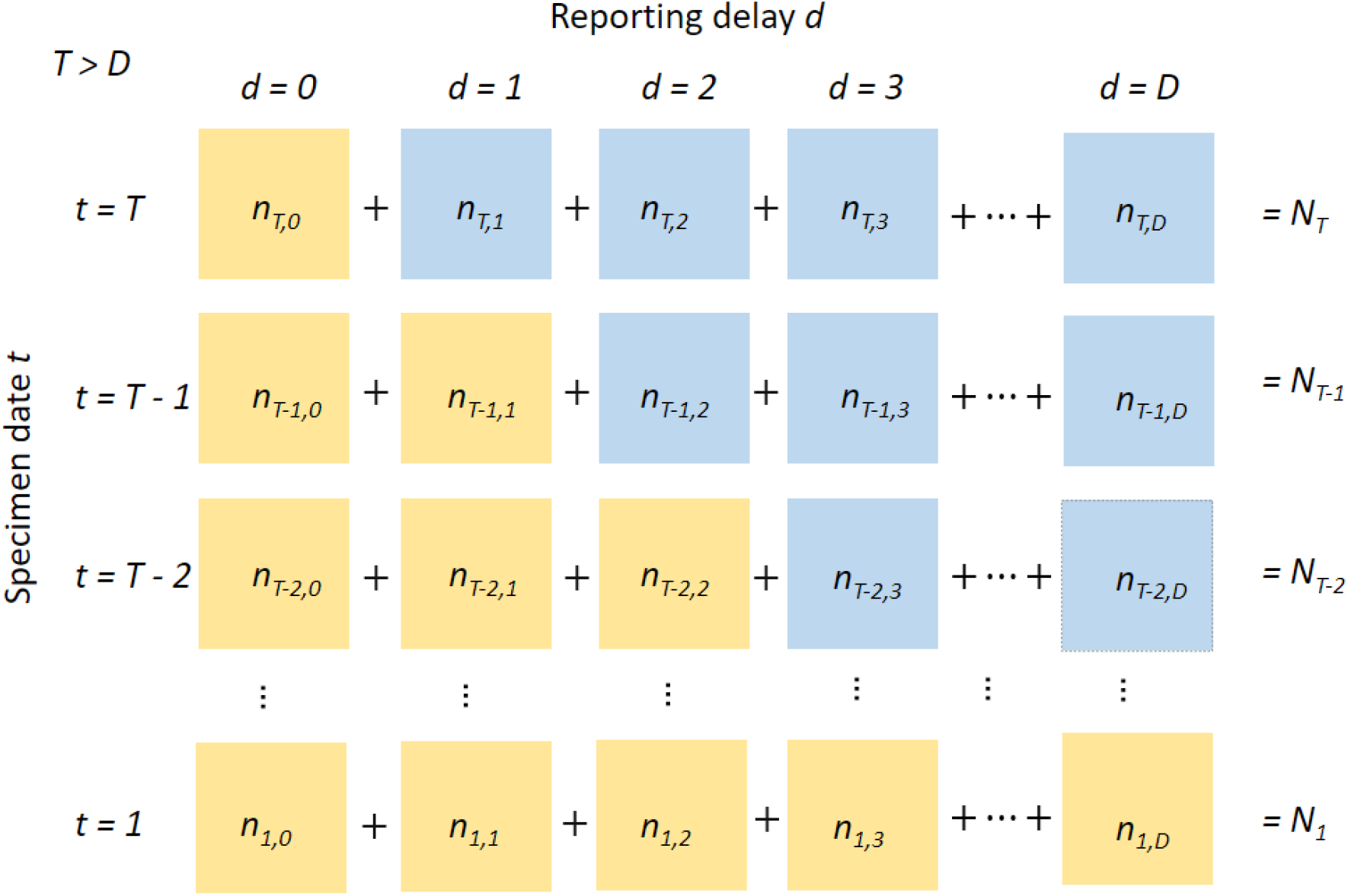
Illustration of the reporting delay data triangle structure, with elements of the 2-dimensional array. Horizontal axis represents the report delay and vertical axis the specimen date. Complete data per specimen date correspond to the sum of each row across the reporting delays. Each cell represents the case count for a given specimen date and reporting delay. Case counts are unknown in real-time when d > T-t, represented here by blue cells.

The element *n*_*t,d*_ represents the number of tests collected on the *t*^th^ day of the specimen date time series that were reported after *d* days. In theory,*D* could be very large. However, in practice most reporting delays are under 10 days. Therefore, for this analysis, we assume a maximum possible reported delay of 50, though each model may assume a shorter value. In real-time, cases *n*_*t,d*_ cannot exist when *d* >*T* −*t* which introduces a right truncation.

Therefore, for cases at *t* = *T* only cases with *d* = 0 can be known, with other values 1 ≤ *d* ≤*D* unknown. The quantity of most interest used to inform decision making and proactive communications was the total cases by specimen date *N*_*t*_. The reporting triangle is therefore collapsed into 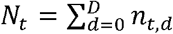. To support operational needs, these daily counts are also aggregated to weekly levels for ease of interpretation.

#### NHS 111 Online Pathways

While there is a delay in case reporting, other data sources are complete in real-time and rapidly available. These data were be leveraged to inform case prediction. NHS 111 Online Pathways is an online triage service in the UK used to give non-emergency healthcare guidance to individuals [17]. Users are routed to appropriate guidance given input information about their symptoms. We transformed these inputs into symptom categories,, and calculate counts of symptom triages,, by time, and symptom category. Symptom categories and groupings are given in Supplementary Table 1, with visualisations of the trends in Supplementary Figure 2 & 3.

### Models

The aim of our nowcasting models was to estimate the expected complete number of cases that have been collected during the most recent 7 days,. Some models harness the partial reporting of recent cases correcting for the delay distribution, others ignore this partial reporting. We aimed to select methods that perform well against the norovirus dynamics observed. Models were tuned for appropriate parameter selection over the 4-week period using weeks ending 8 October 2023 to 29 October 2023, then applied to the remainder of the weeks to 10 March 2024, to avoid parameter selection using evaluation data. Models are tuned based on the average daily scores for the most recent 7 days, as outlined in the evaluation section. Model structures and assumptions are given in Table 1.

**Table 1.** Summary of key model structures, assumptions, and characteristics to compare for each model.

| Property | Baseline | BSTS | BSTS + NHS 111 online | GAM | epinowcast |
| --- | --- | --- | --- | --- | --- |
| Uses partial reported data | No | No | No | Yes | Yes |
| Parametric reporting delay distribution | - | - | - | No | Yes |
| Supplementary indicator signal | No | No | Yes | No | No |
| Bayesian | No | Yes | Yes | No | Yes |
| Parameter estimation method | - | Gibbs | Gibbs | REML, sampling via Metropolis-Hastings | HMC with NUTS |
| Approximate runtime per week (fitting and post-processing) | 0.1s | 50s | 1 min 45s | 10s | 10 min |
| Posterior samples (burn-in) | - | 50,000 (2,000) | 50,000 (2,000) | 1,000 (1,000) | 1,000 (1,000) |

#### Baseline

To contextualise the performance of the models, we implemented a simple baseline approach to compare against. We assumed each predicted day will be equal to the observed count the previous week giving an autocorrelated prediction with day-of-week effects.

The central estimate is set as 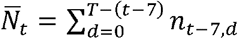, which corresponds to the reported data from the seven days prior – matching the weekly reporting cycle in surveillance. Most norovirus cases were reported with *d* ≤7 and as such this method gives predictions of near complete case numbers. We did not consider uncertainty within the baseline method. For application of the scoring methodology, prediction intervals are required. Therefore, for the baseline model the prediction intervals were assumed equal to the central estimate.

#### Generalised Additive Model

We used a generalised additive model (GAM) utilising partially reported data, based on a nowcasting model for mpox [18] [19]. This estimated the total number of cases with specimen date 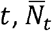 as the sum of known data that has already been reported, *n*_*t,d*_, for reporting delays *d* ∈ [0,*T*−*t*], and estimates for the unknown data yet to be reported,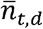, for reporting delays *d* ∈ [*T*−*t* +1,*D*],i.e.

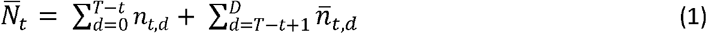

As *n*_*t,d*_ is known, 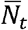 has a natural lower bound of 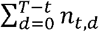. The unknown data was modelled with a negative binomial distribution accounting for the non-negative integer values and overdispersion. Using the mean and variance parameterisation,

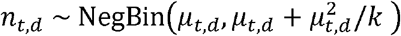

with dispersion parameter *k*. We use a log link function to model the exponential epidemi process, where *μ* _*t,d*_ depends on both *t* and *d* according to

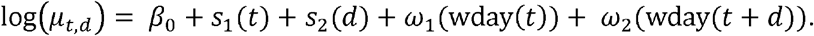

where *β*_0_ is a constant. We assumed that the number of cases vary smoothly over specimen date *t* and number of days delay *d* as *s*_1_(*t*) and *s*_2_(*d*), with random day-of-week effects *ω*_1_(wday(*t*)) and *ω*_2_ (wday(*t*+*d*)) respectively. The model was fitted in *R* using the *gam* from the *mgcv* package [20]. 1000 burn-in and posterior samples were drawn from for the model using the *gratia* package [21] with a Metropolis-Hastings sampler. Samples were aggregated to 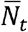 (eqn. 1), with prediction intervals taken using quantiles of these samples. Models were fit to the past 56 days, with cubic regression basis functions every *l*=7 days for *s*_1_(*t*) and *s*_2_(*d*), and a maximum reporting delay *D* = 14. Model tuning is outlined in Supplementary Section 2.

#### Epinowcast

We also used a Bayesian hierarchical nowcasting framework implemented in the *epinowcast* package [22], with the implementation described below. This approach builds on earlier nowcasting approaches [23] [24]. As with the “GAM” model, the estimate for the total number of tests with specimen date 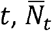, is the sum of known data, *n*_*t,d*_, and estimates for the unknown data, 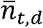 (eqn. 1).

Here *n*_*t,d*_ |*N*_*t*_ follows a multinomial distribution with a probability vector (*P*_*t,d*_) that is estimated jointly with the expected number of final reported cases. This differs from the “GAM” model approach, where each *n*_*t,d*_ is independent. We used the default implementation of modelling expected final reported cases as a first order random walk,

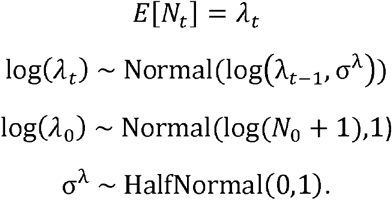

The instantaneous growth rate *r*_*t*_ is defined as the log of the expected number of final reported tests between time *t* and *t* − 1.*r*_*t*_ is then modelled on the log scale by a daily random effect *ω*_1_(*t*) and a random effect for the day of the week *ω*_2_(wday(*t*)),to account for weekly periodicity in the underlying data.

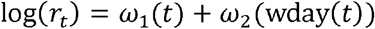

Within *epinowcast* the delay distribution (*p*_*t,d*_) is then defined as a discrete time hazard model where:

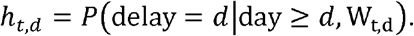

Here, the hazard is determined by a design matrix W_t,d_ including a baseline delay distribution and time- and delay-specific covariates which affect the reporting delay. We assume the probability of reporting 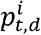 follows a discretised log-normal distribution where the log mean and log standard deviation are modelled with a daily random effect (the model default).

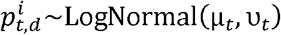

where the parametric logit hazard γ_*td*_ is given by

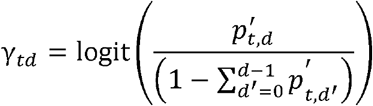

We also use a constant non-parametric logit hazard such that:

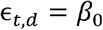

The overall hazard is then modelled as logit (*h*_*t,d*_)= γ _*t,d*_ + ∈_*t,d*_

To estimate final observed reported cases a negative binomial observation model is used where:

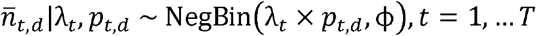

and the overdispersion parameter ϕ is estimated with a prior of

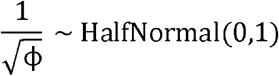

and 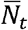 is given by (eqn. 1).

Unlike the “GAM” model, this approach introduces parametric, discrete, and truncated distributions for the reporting delay, better reflecting the reporting measurements. Models are fit in *stan* with *cmdstan* [25] using the Hamiltonian Monte Carlo (HMC) with NUTS (No-U-Turn Sampler). We ran 1000 iterations for warm-up and 1000 post-warmup iterations. A maximum reporting delay of 7 days, with a training length of 35 was selected. Model tuning and prior specification are outlined in Supplementary Section 3.

#### Bayesian Structural Time Series

We employed a flexible Bayesian structural time series (BSTS) modelling approach to produce a nowcast without harnessing partial reported case counts. The time series *N*_*t*_ is truncated by 7 days, with the unknown daily counts estimated in a traditional forecasting approach. The BSTS allows for a state space specification with decomposition of time varying dynamics including trend, seasonality and regression effects [26]. We create two models using the *bsts* R package [27], one without regressors, the second using 111 online indicators.

The first model “BSTS” is defined by the following state space equations, where at time *t*, we have mean *μ*_*t*_, slope *δ*_*t*_ and seasonal component *τ*_*t*_, with a season as *S* = 7 days to capture the day-of-week effects.

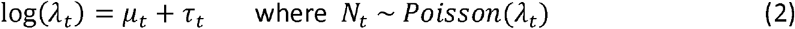

The equation for the mean *μ*_*t*_ is given by

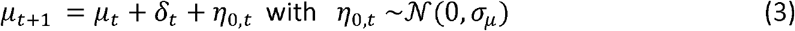

and the slope,

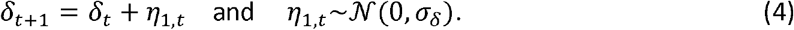

Lastly the seasonality component is determined via dummy regression variables,

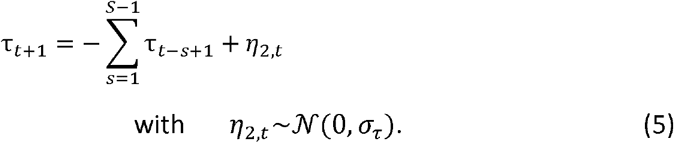

This ensures that the seasonal component τ_*t*_ accounts for the cumulative seasonal effects over the specified period *S*, in our case one week. Therefore, log(*λ*_*t*_) follows a local linear trend with seasonality, where the mean and slope of the trend are assumed to follow random walks. For the “BSTS” model, a training length of 60 days was chosen, with upper limits of exp(*σ* _*μ*_) and exp(*σ* _*δ*_) equal to 1.1. Model tuning is outlined in Supplementary Section 4. The models were fit via Gibbs sampling MCMC, run for 50,000 iterations with 2,000 burn in.

To produce the second model “BSTS + NHS 111 online” we update the observational equation (1) to include the regressor symptom category scaled counts *x*_*i,t*_ in ***x***_*t*_

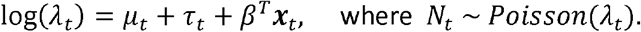

The *β*_*i*_ values are estimated using spike and slab priors [28] centred on zero to allow for sensible variable selection. For the “BSTS + NHS 111 online” model we choose a training length of 150 days, 5 expected regression coefficients (through the spike and slab prior), and an upper limit for exp(*σ*_*µ*_) of 1.01 and exp(*σ* _*δ*_) of 1.1. Model tuning analysis is given in Supplementary Section 4.

### Models Overview

### Evaluation

To compare the different nowcasting approaches we employ multiple scoring methods in a probabilistic framework. The interval coverage is a measure of probabilistic calibration, telling us the proportion of observations that are within given prediction interval ranges – in our case 50% and 90%. From the interval coverage we calculate the coverage deviation, the average difference between the measured interval coverage and the specified interval value, with a coverage deviation nearer zero being preferred. The (weighted) interval score (WIS) is a proper scoring rule composed of sharpness and under/overprediction, giving an overall measure of performance where low values are better. The weighted interval skill score is calculated as 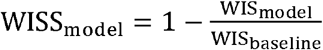 where WISS_model_ > 0 corresponds to a model better than the “baseline” model. The bias is a relative measure of under/overprediction telling us if the models systematically estimate high or low, with lower bias models having a score nearer zero. The median absolute error gives an average of the absolute difference between central prediction and true data. The scoring is conducted using the *scoringutils* package [29]. The estimates are scored at daily and weekly aggregations, as well as explored by nowcast horizon *h*,where *h* = *T* −*t* in our case is the day-of-week predicted. Since the data is uploaded weekly, the nowcast horizon *h* corresponds to a unique day-of-week where Sunday will be a nowcast horizon of 0 days, and *Monday* will have a nowcast horizon of 6 days.

## Results

Winter 2023/2024 followed the seasonal trend of increasing cases from September onwards, reaching a stable trend from December 2023 onwards. The difference between final and initial cases is largest in the most recent days each week, as expected, with *n*_*t*,0_ near zero (Figure 2a). Across each week approximately 20% of the data are revisions (cases added the following week). These revisions can change the narrative of the real-time trend without correction (Figure 2b). The distribution of shows few reports on *d* = 0, a peak at 1-2 days and most reports within 7 days (Figure 3). The time varying reporting delay is given in Supplementary Figure 4, showing limited variation.

**Figure 2.**
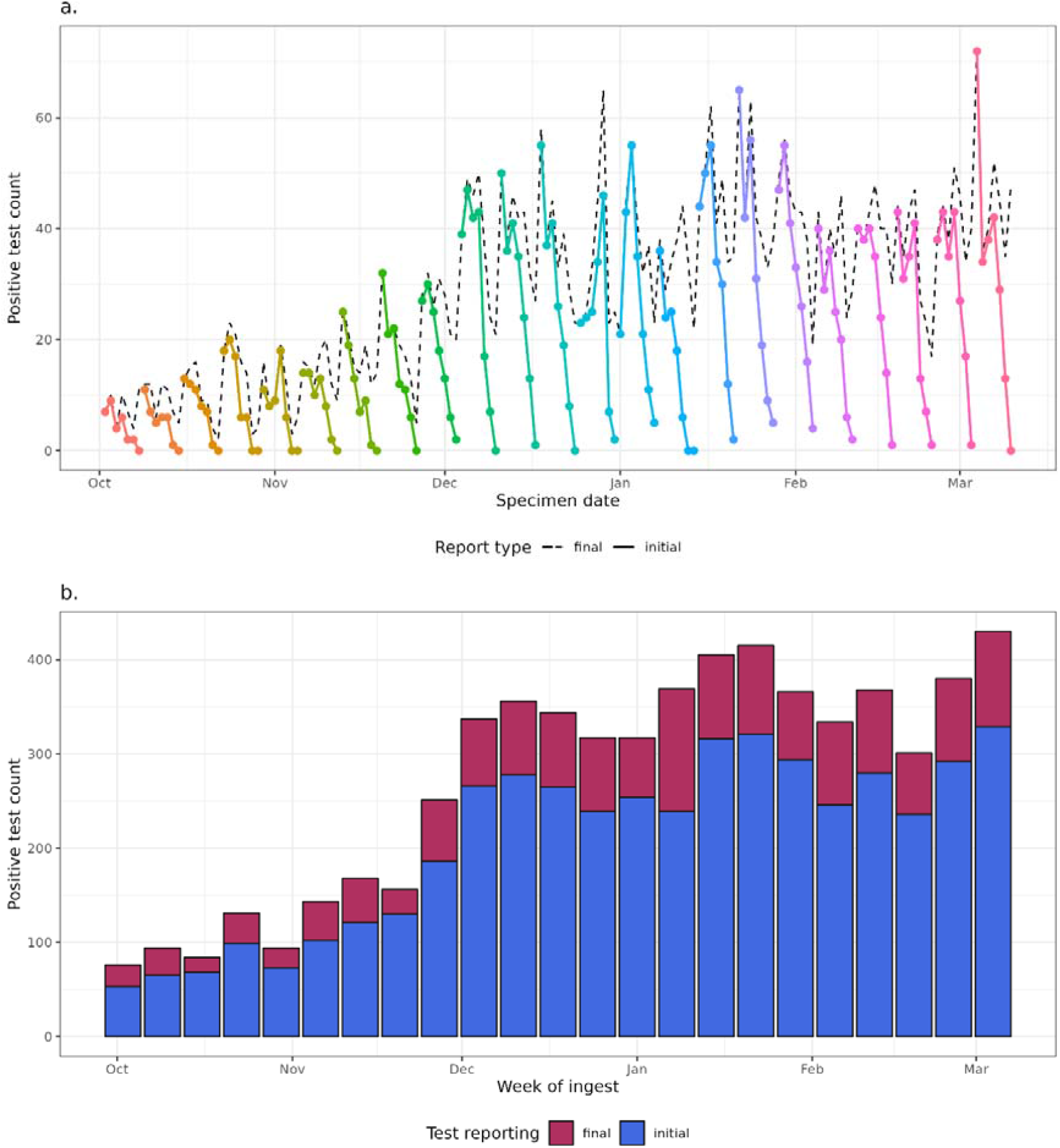
The backfilling of norovirus tests over the Winter 2023/2024 season. (a.) daily counts of tests at different snapshots of reporting, showing the most recent observed counts are substantially lower than the final revised data. (b.) weekly counts of tests at each ingest and final revisions. The end date for each week was taken as a Sunday, to produce a nowcast of data from the previous week.

**Figure 3.**
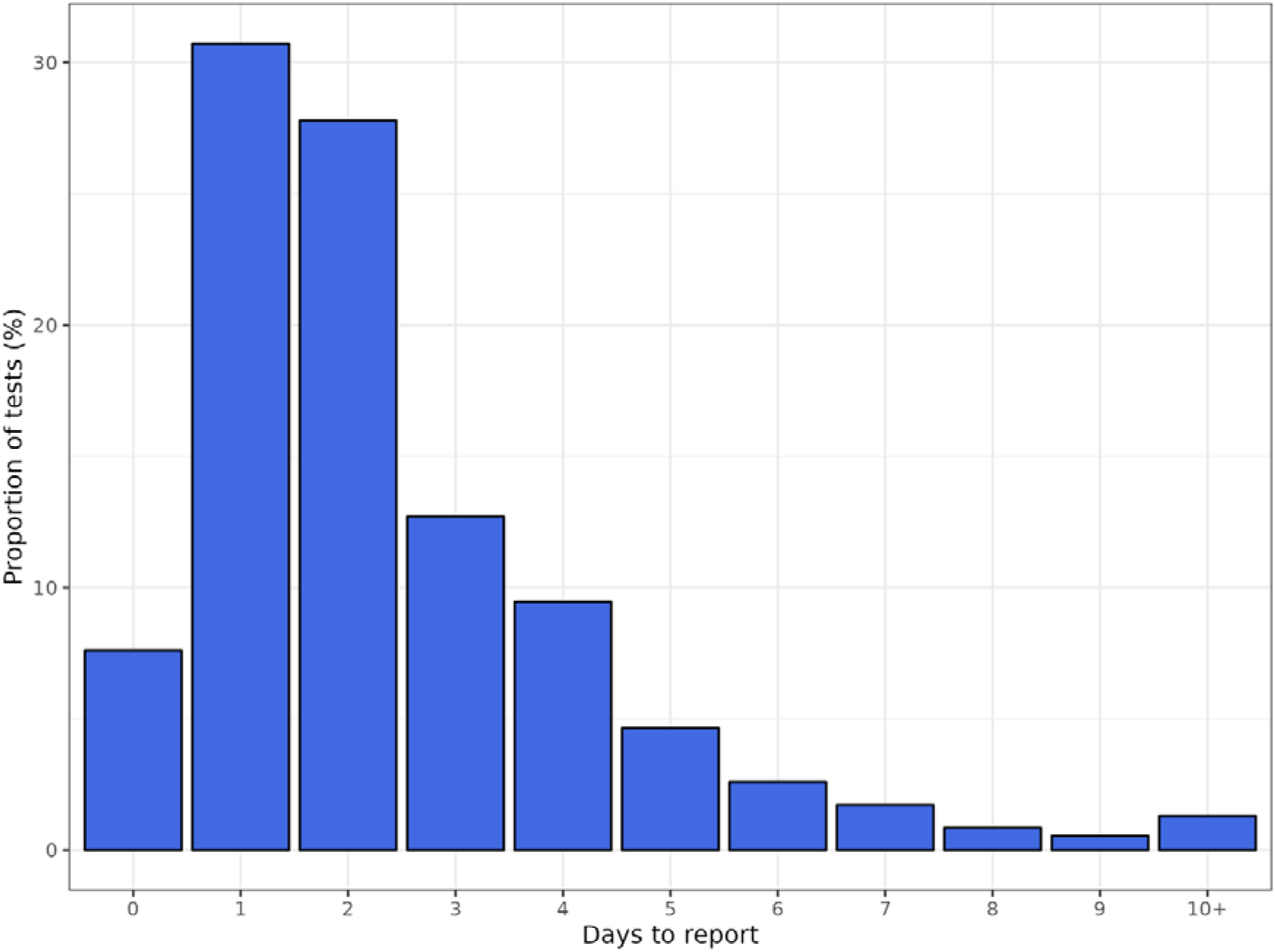
Time delay distribution of days between specimen date and report date. Includes complete data from 02-10-2023 to 10-03-2024.

The daily and weekly nowcasts are shown over the tuning and evaluation time periods (Figure 4 & 5). Both the “GAM” and “epinowcast” models show increasing uncertainty towards the most recent date where data is more incomplete. The models using the partially complete data underpredict the complete cases in the week ending 14 January 2024, which we also see in the weekly estimates (Figure 5), though the “BSTS” is not impacted in this way. The uncertainty in the weekly estimate varies substantially by model, though the “baseline” model has no associated uncertainty. The BSTS models have wide prediction intervals compared to the “GAM”, with the “epinowcast” model prediction intervals being skewed towards higher values.

**Figure 4.**
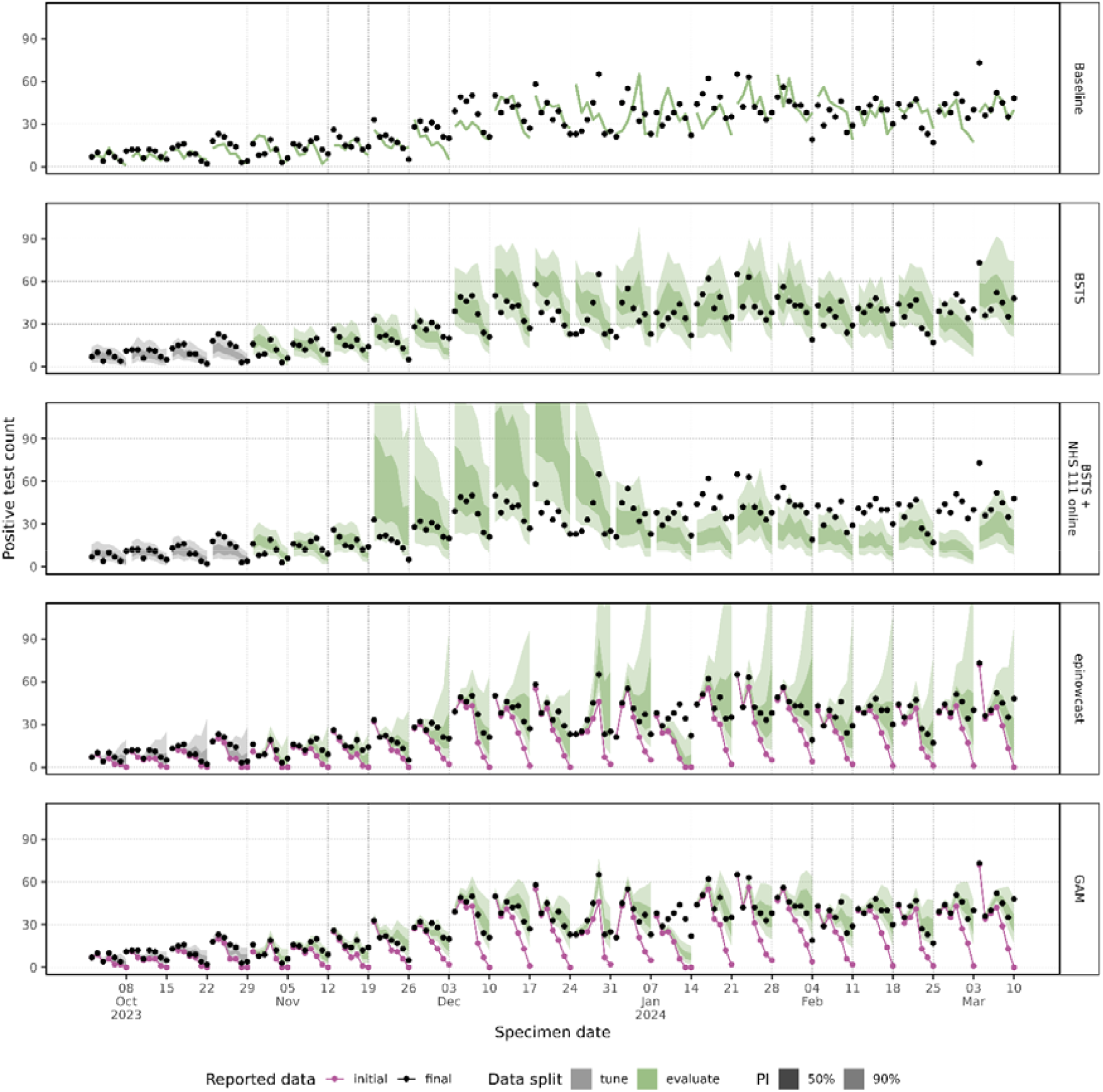
Daily predictions from all models with 50% and 90% prediction intervals against initial and final reported count of tests.

**Figure 5.**
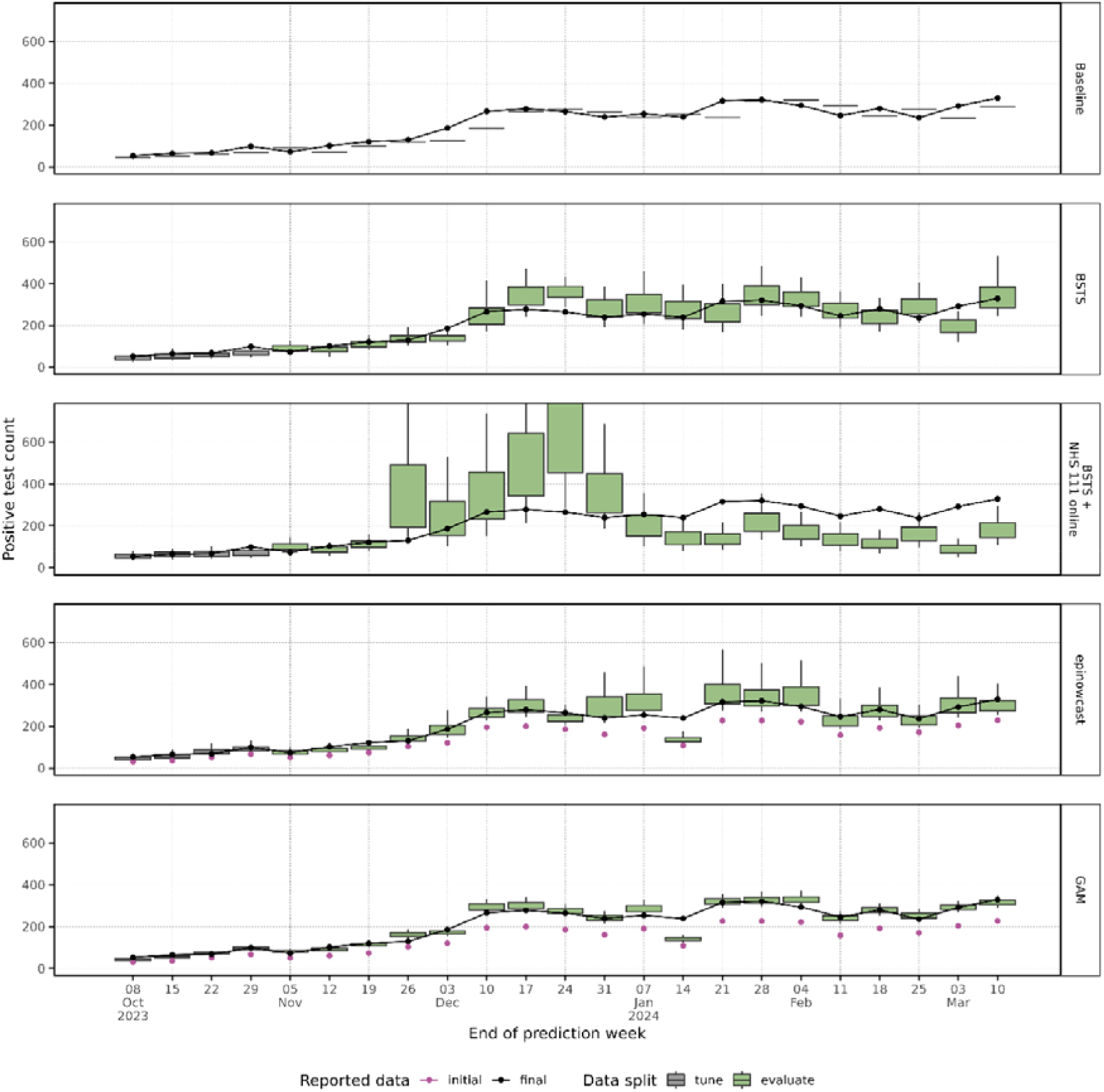
Weekly predictions from all models with 50% (box) and 90% (whiskers) prediction intervals against initial and final reported count of tests. The weekly predictions are created as the sum of sample predictions per week.

The overall daily and weekly evaluation scores are shown in Table 2. The “baseline” model has high WIS, expected given its small interval width. The partial reporting delay models “epinowcast” and “GAM” outperform other models across WIS and MAE, generally overpredicting, when other models are underpredicting. The “BSTS” model performs better than the baseline across all daily metrics, whereas the “BSTS + NHS 111 online” performs broadly worst. Across daily and weekly scoring the “BSTS” model has the best calibration with lowest coverage deviation, though other models have similar values. Notably, the “GAM” and “epinowcast” models over and underpredict respectively.

**Table 2.** Breakdown of overall model scores by temporal granularity. The daily granularity shows the average daily score over the time series. The weekly granularity shows the average weekly score over the time series. The most optimal score by temporal granularity and scoring metric is in bold.

| Temporal granularity | Model | WIS | Median absolute error | Bias | Coverage deviation |
| --- | --- | --- | --- | --- | --- |
| daily | Baseline | 7.73 | 7.73 | -0.21 | -0.49 |
| daily | BSTS | 4.57 | 7.18 | -0.07 | **0.03** |
| daily | BSTS + NHS 111 online | 10.28 | 15.36 | -0.24 | -0.12 |
| daily | epinowcast | 3.03 | 4.07 | -0.31 | 0.08 |
| daily | GAM | **2.29** | **3.39** | **0.05** | 0.05 |
| weekly | Baseline | 29.74 | 29.74 | -0.39 | -0.56 |
| weekly | BSTS | 21.19 | 34.04 | -0.05 | **-0.06** |
| weekly | BSTS + NHS 111 online | 67.44 | 100.35 | -0.30 | -0.20 |
| weekly | epinowcast | 15.61 | 22.35 | -0.19 | 0.08 |
| weekly | GAM | **11.56** | **16.00** | **0.04** | -0.11 |

Over the evaluation period the “GAM”, “BSTS” and “epinowcast” models have improved skill over the baseline model in most but not all weeks (Figure 6c). For much of the time series, the “BSTS+NHS 111 online” model has higher WIS than the baseline model (Figure 6b). The “GAM” and “epinowcast” models have bias > 0 during the epidemic growth phase, indicating overprediction (Figure 6c). The week of 14 January 2024 the “epinowcast” and “GAM” perform markedly worse than other weeks, where initial reported data is particularly low. Further scoring at daily and weekly levels are given in Supplementary Figures 5 & 6.

**Figure 6.**
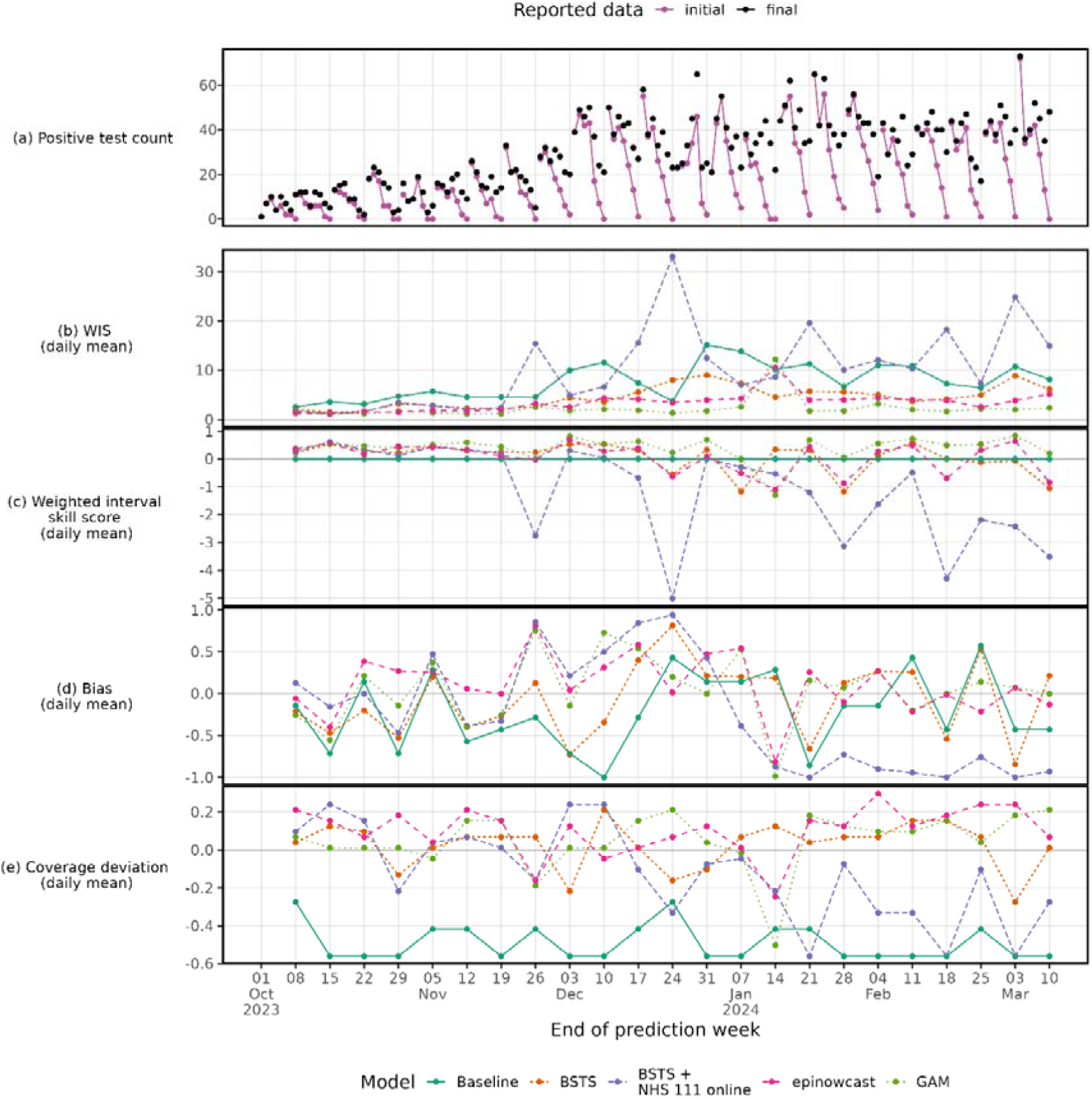
Daily count of final and initial reported tests (a) with daily mean model scores for each prediction week. The Weighted Interval Score (b), Weighted Interval Skill Score (c), Bias (d) and Coverage deviation (e) are given across models and time.

By breaking down by the day-of-week (and therefore nowcast horizon, in our case) we can explore how varying data completeness affects model performance. Relative to “baseline” the “BSTS” model exhibits a flat skill across days (Figure 7a), whereas the relative skill of the “GAM” and “epinowcast” gets deteriorates towards the end of the week (Figure 7b). The “baseline” consistently underpredicts, while “epinowcast” underpredicts at the start of the week but becomes less biased toward Sunday (Figure 7c). Compared to the “BSTS” model, the improved performance of the “GAM” model is primarily due to lower WIS early in the prediction week when data is more complete.

**Figure 7.**
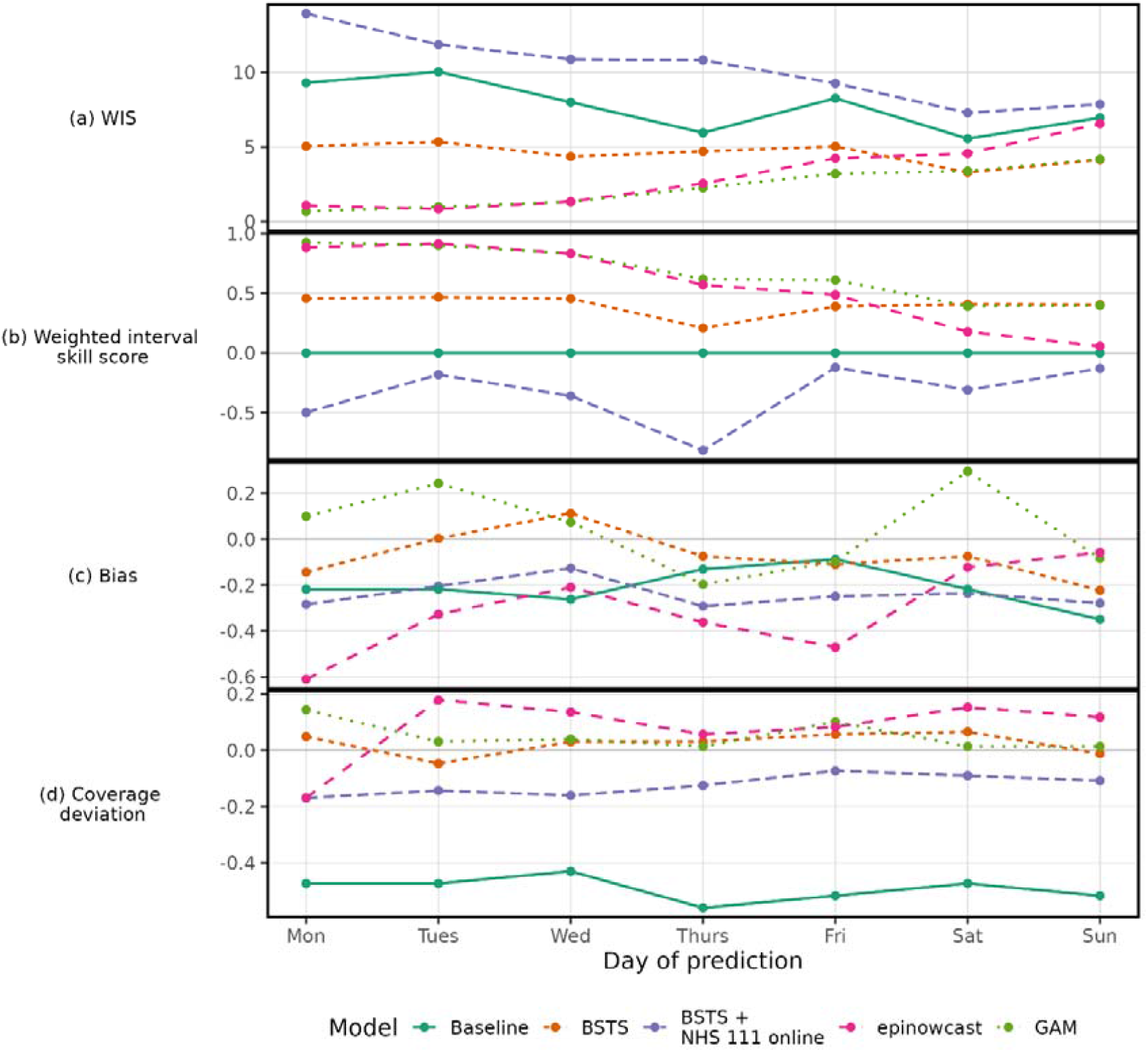
Model scores averaged over each day of prediction. A Monday has near complete data, whereas a Sunday has many cases not yet reported. The scores are the average over the evaluation period.

## Discussion

Norovirus contributes substantially to health service winter pressures through hospital outbreaks, reduced bed availability and staff absences. As such, timely surveillance is crucial for situational awareness, particularly to understand changes in the epidemic curve in the context of delayed reporting. In this work we applied a range of nowcasting approaches to norovirus cases, with the aim of understanding the current epidemic state.. We have shown that harnessing partially complete data outperforms a truncate-and-forecast approach, but the performance can be sensitive to the consistency of case reporting, which is challenging in frontline health protection. The delay in reporting impacts the analysis of trends in national surveillance, so it is important official reporting exclude these partially reported days, though nowcasting can support decision making in real-time. The nowcasting problem presented is a straightforward application of time delay correction, with a small average delay, a single test type, and without considering regional or age-related variation. This may partially explain the strong performance of approximate methods in the scoring.

Nowcasting approaches are increasingly used to predict case counts by accounting for delays in reporting, and have been crucial in the recent COVID-19 pandemic and mpox outbreak [12] [24] [18] [30]. In this analysis, we apply several modelling approaches from the epidemic literature to this problem. We compare a well-principled Bayesian implementation, *epinowcast*, which jointly models a reporting delay distribution with an underlying process model, and a more approximate but highly flexible and computationally efficient GAM-based model. We also consider a Bayesian structural time series approach, testing the utility of incorporating leading indicators into the modelling framework. To our knowledge this is the first study to apply time delay nowcasting methods to norovirus cases, which may be more challenging to nowcast than other infectious diseases due to high levels of underreporting, regional heterogeneity and its association with outbreaks in closed settings such as care homes, schools and hospitals [31]. Despite this, several models generated operationally useful predictions of norovirus test counts, offering a substantial improvement over using truncated data (the current standard) or a naïve seasonal baseline. However, when reporting delay data is unavailable, time series forecasting presents an adaptive alternative with good coverage and performance compared to the baseline. In contrast to previous studies, we did not find including leading indicators improved our predictions [32]. This could perhaps be explained by lower signal in the indicators considered, related to confounding effects from other winter pathogens. Finally, our findings that several models perform well with different accuracy and biases over time and day of the week suggests the potential benefit of an ensemble approach, as has been demonstrated in other contexts [12].

Models incorporating reporting delays consistently performed better than forecasting models that do not, showing the utility of leveraging this data when available. This improved performance is driven by reduced uncertainty when there is more complete reported data, early in the nowcast window. Among our models using reporting delays, we found that the time delay approximation method in the “GAM” scored slightly better than the more complex “epinowcast” model’s full joint distribution approach, in this application. The “epinowcast” has increased uncertainty due to modelling the reporting delay distribution and underlying process model. Wide intervals are penalised in scoring metrics like the WIS, however, this larger uncertainty may better reflect the uncertainty in the system. We saw that modelling based on recent distributions of reporting delays can perform poorly if these distributions change rapidly, although in these cases, the “epinowcast” model’s optional time-varying delay may be advantageous compared to a fixed distribution approach, such as the one in the “GAM”. Speed is key in a real-time modelling context, with some models being substantially faster than others, however, all approaches ran in a reasonable time (Table 1) for real-time inference. The computational expense of “epinowcast” compared to other models, however, was impactful during model development.

The performance of some models may have been limited due to the tuning approach taken. Hyperparameter optimisation was performed on a time before the epidemic wave started, simulating a plausible real-time scenario – which may bias selection toward hyperparameters good at flat periods of incidence. There are reporting changes in frontline healthcare delivery which can impact the performance of time delay informed models – these local practices are challenging to understand in real-time and adjust for in modelling, which should be explored further. Future work should explore how local testing practices can be incorporated into modelling directly. Understanding testing pathways and real-time modelling of norovirus will be crucial for the next strain replacement event highlighting the importance of developing our understanding and preparedness.

While not a high priority pandemic potential pathogen, norovirus causes healthcare system strain and an unpleasant infection for the individual, increasing associated opportunity cost by blocking beds and elongating patient length of stay [3]. Estimating the current case burden when accounting for delayed reporting can be an important tool for supporting effective public health response. In this work we have compared the options available to correct for delayed reporting, highlighting their strengths and limitations – notably demonstrating the importance of explicitly modelling the partially complete data. This work will underpin situational awareness should the next strain replacement event occur.

## Data Availability

Training data for the models explored in this manuscript is available at https://github.com/jonathonmellor/norovirus-nowcast. This data is aggregate with statistical noise added to preserve anonymity. This data enables each model to be fit and can be used for the future development of nowcasting models. Code for running all models is available at https://github.com/jonathonmellor/norovirus-nowcast. Individual-level data on the reporting delay used to inform initial exploration are not available due to patient identifiability. An application for data access can be make to the UK Health Security Agency.
UKHSA operates a robust governance process for applying to access protected data that considers:
- the benefits and risks of how the data will be used
- compliance with policy, regulatory and ethical obligations
- data minimisation
- how the confidentiality, integrity, and availability will be maintained
- retention, archival, and disposal requirements
- best practice for protecting data, including the application of 'privacy by design and by default', emerging privacy conserving technologies and contractual controls
Access to protected data is always strictly controlled using legally binding data sharing contracts.
UKHSA welcomes data applications from organisations looking to use protected data for public health purposes.
To request an application pack or discuss a request for UKHSA data you would like to submit, contact.

https://github.com/jonathonmellor/norovirus-nowcast

## Contributions

**JM** – Conceptualisation, Methodology, Software, Validation, Formal Analysis, Data Curation, Writing – Original Draft, Writing – Review & Editing, Visualisation, Project Administration

**MT** - Methodology, Software, Validation, Formal Analysis, Data Curation, Writing – Original Draft, Writing – Review & Editing, Visualisation

**EF** - Methodology, Software, Formal Analysis, Writing – Original Draft, Writing – Review & Editing, Visualisation

**RC** – Conceptualisation, Data Curation, Writing – Review and Editing

**OP** - Software, Formal Analysis, Writing – Review & Editing

**CEO** – Methodology, Writing – Review & Editing

**AH** – Investigation, Writing – Review & Editing

**AD** – Conceptualisation, Investigation, Writing – Review & Editing

**SRD** – Conceptualisation, Writing – Review & Editing, Supervision

**TW** – Writing – Review & Editing, Supervision

## Ethical Approval

UKHSA have an exemption under regulation 3 of section 251 of the National Health Service Act (2006) to allow identifiable patient information to be processed to diagnose, control, prevent, or recognise trends in, communicable diseases and other risks to public health.

## Data Availability Statement

Training data for the models explored in this manuscript is available at https://github.com/jonathonmellor/norovirus-nowcast. This data is aggregate with statistical noise added to preserve anonymity. This data enables each model to be fit and can be used for the future development of nowcasting models. Code for running all models is available at https://github.com/jonathonmellor/norovirus-nowcast. Individual-level data on the reporting delay used to inform initial exploration are not available due to patient identifiability. An application for data access can be make to the UK Health Security Agency. UKHSA⍰operates a robust governance process for applying to access protected data that considers:

- the benefits and risks of how the data will be used
- compliance with policy, regulatory and ethical obligations
- data minimisation
- how the confidentiality, integrity, and availability will be maintained
- retention, archival, and disposal requirements
- best practice for protecting data, including the application of ‘privacy by design and by default’, emerging privacy conserving technologies and contractual controls

Access to protected data is always strictly controlled using legally binding data sharing contracts.

UKHSA⍰welcomes data applications from organisations looking to use protected data for public health purposes.

To request an application pack or discuss a request for UKHSA data you would like to submit, contact.

