## Supplementary Information for "An Application of Nowcasting Methods: Cases of Norovirus during the Winter 2023/2024 in England"

**Section 1**

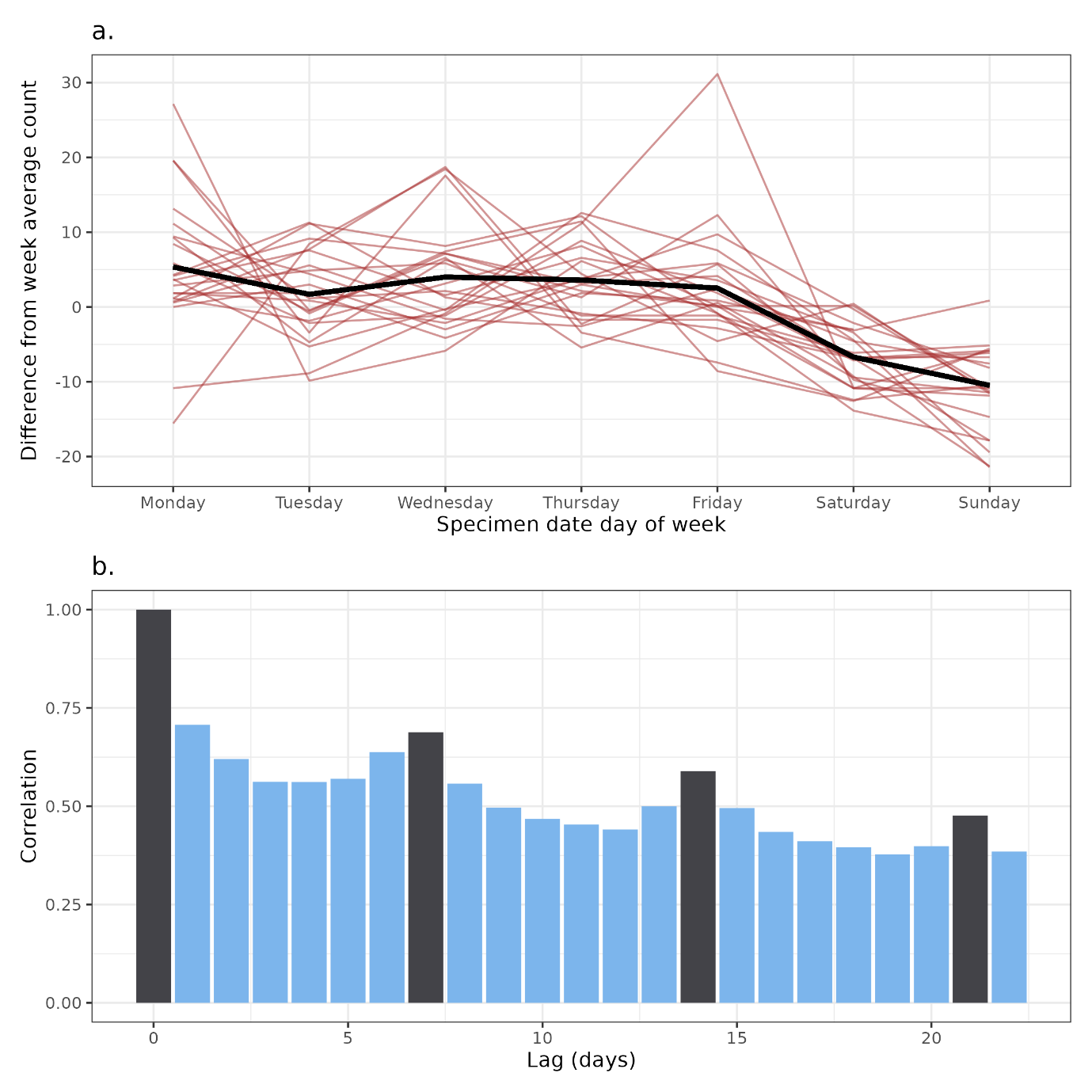

Supplementary Figure 1. Analysis of the periodicity (a.) and autocorrelation (b.) of norovirus aggregate counts by specimen date. (a.) shows the difference between the count of tests on a given day and the average of counts that week (red), with the mean value per day (black). (b.) shows the autocorrelation between counts of positive tests with a clear weekly periodicity shown by high values each 7 days.

| **Indicator name** | **Condition – pathway description** |
| --- | --- |
| gastro intestinal (GI) symptoms | Contains “diarrhoea” or “vomit” or “nausea” |
| fever | Contains “fever” or “high temperature” |
| headache | Contains “headache” or “migraine” |
| limb pain | Contains “arm pain” or “painful arm” or “leg pain” or “painful leg” or “limb pain” |
| stomach pain | Contains “abdominal pain” or “stomach ache” |
| not limb or stomach pain | Does not contain “abdominal pain” or “stomach ache” and does not contain “arm pain” or “painful arm” or “leg pain” or “painful leg”, or “limb pain” |

Supplementary Table 1. The logical conditions used to define each indicator from NHS 111 pathways. The texts defining a pathway description change over time, as triage algorithms are updated by NHS 111.

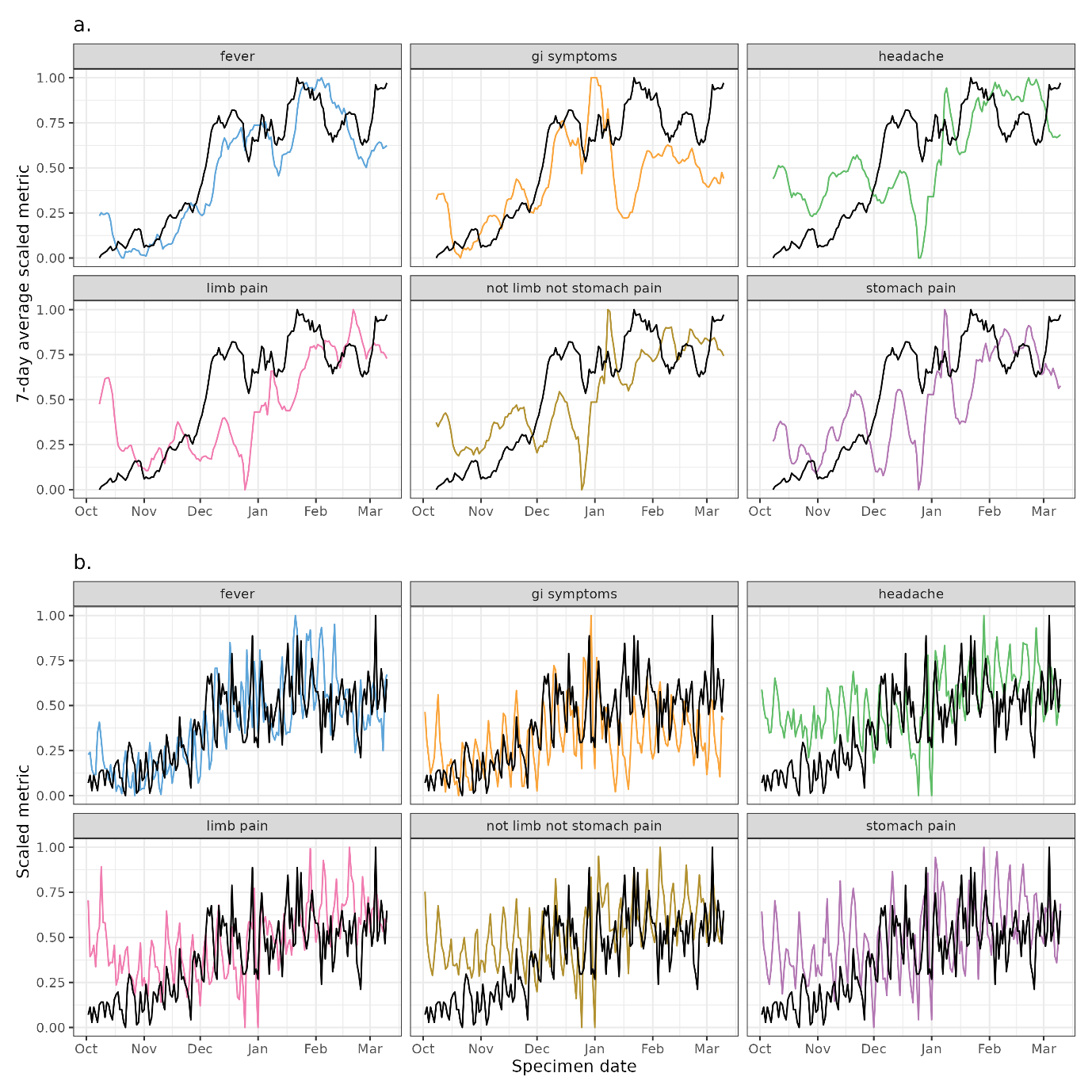

Supplementary Figure 2. The scaled values of different NHS 111 online pathway symptom trends and norovirus positive cases (black). The signals are scaled between 0-1, a.) shows the rolling 7-day mean values for indicator and case trend, where b.) shows the unsmoothed more stochastic data with day-of-week effects.

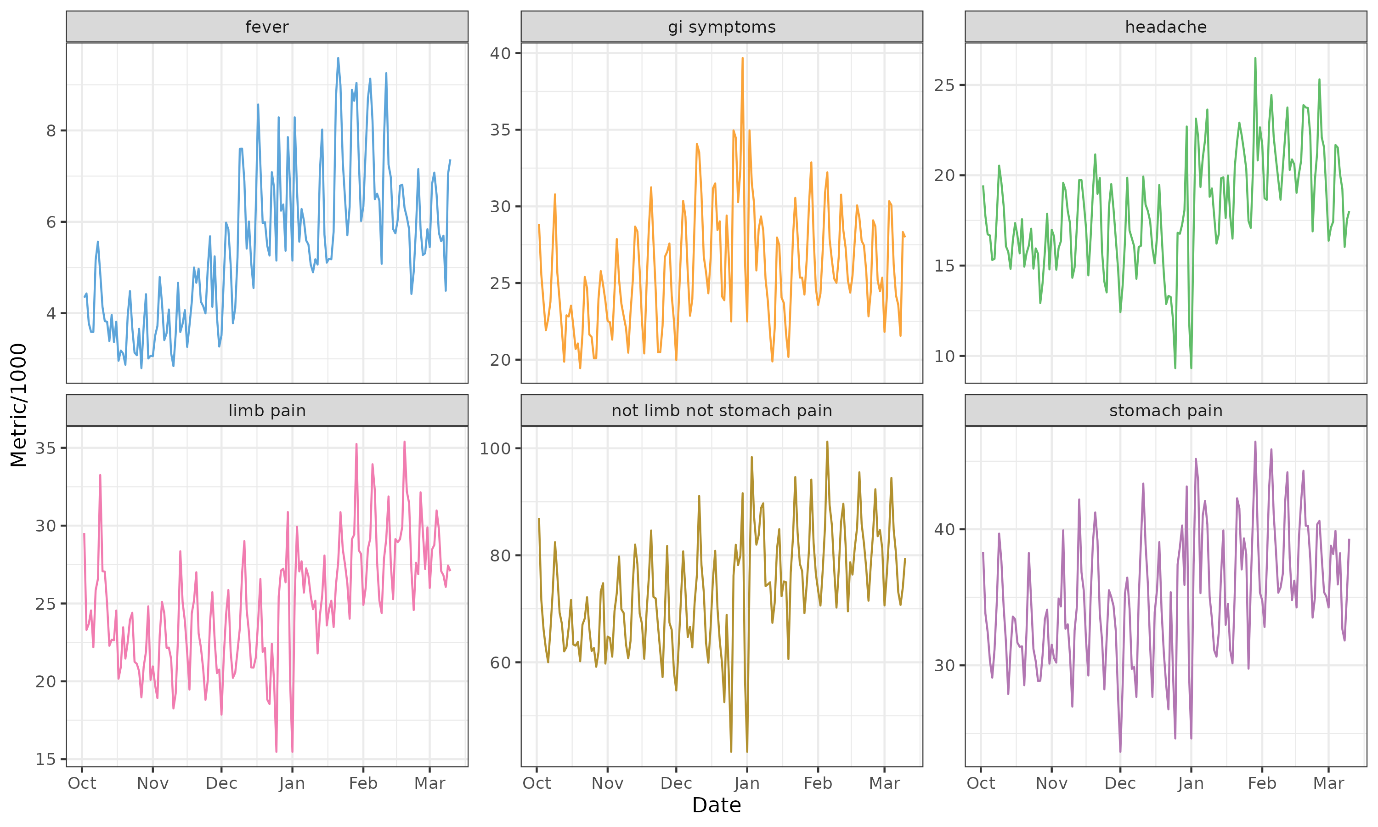

Supplementary Figure 3. Values of each NHS 111 online pathway symptom trends. The more generic symptom categorisations such as “all pain” have larger magnitudes compared to more severe and specific symptoms such as “fever”.

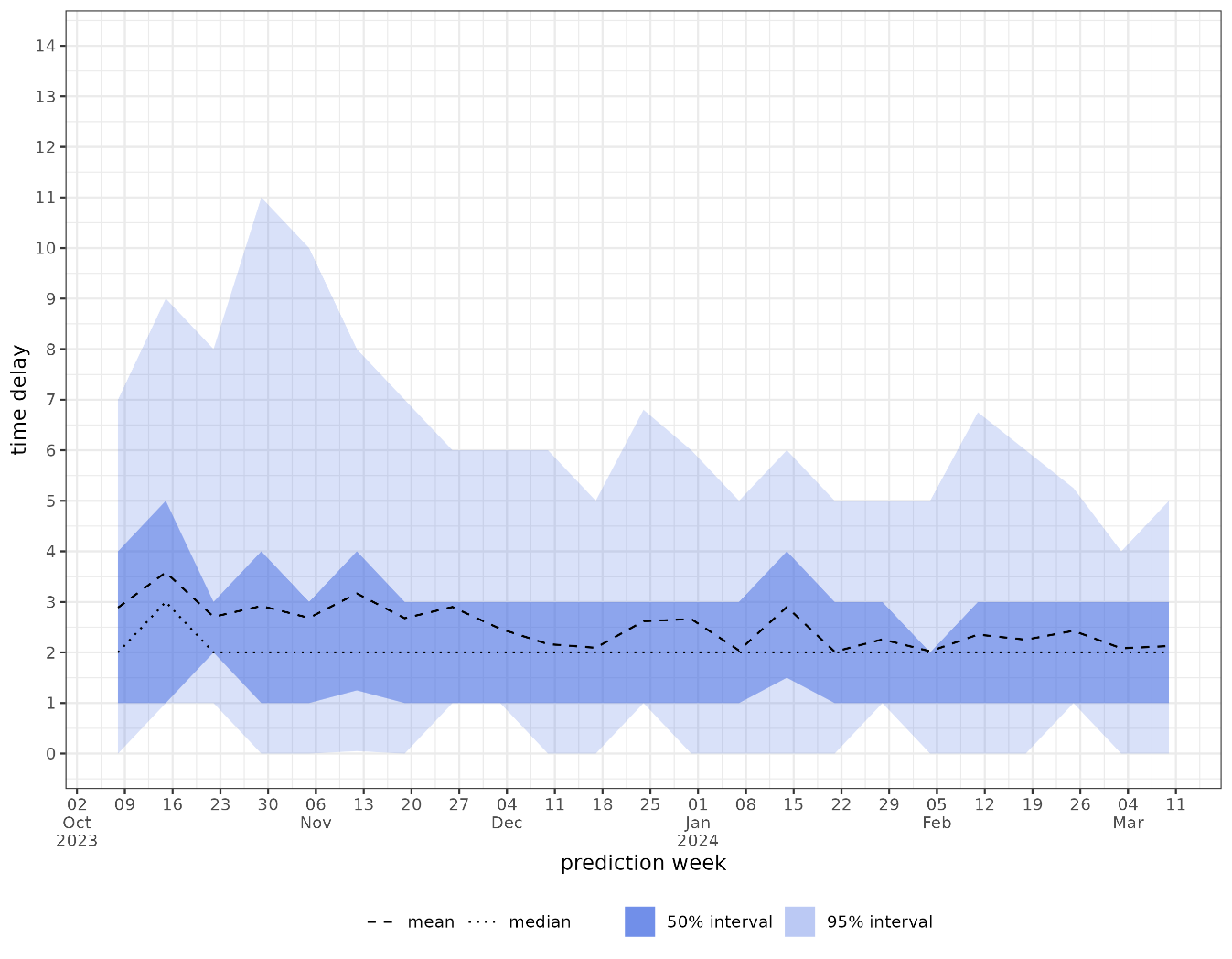

Supplementary Figure 4. The distribution of time from specimen date to report date by nowcast prediction week. The mean, median, 95% and 50% intervals are given for the time delay giving a trend over time. There is a larger tail in reporting delay early in the time series, thought this is the time with fewest positive tests.

**Section 2 – Generalised Additive Model**

Model structure and hyperparameter choices were made to optimise the performance of the model on the most recent 7 days of the nowcast (Supplementary Table 2). For the splines, we use cubic regression basis functions and choose the number of knots every $l$ days. Cubic regression splines were chosen as they performed (via WIS) marginally better than thin plate splines. We consider $l$ as a multiple of 7 to align with the weekly cyclicity, but also evaluate $l=3$ for reporting delay (as values for maximum delay as low as 14 days were tested) – however these models performed worse than those with a larger $l$ for the reporting delay spline. Knots every 7 days for specimen date and 7 days for reporting delay were found to be optimal, along with a training length (relating to specimen date) of 56 days and a maximum reporting delay $D$ of 14 days.

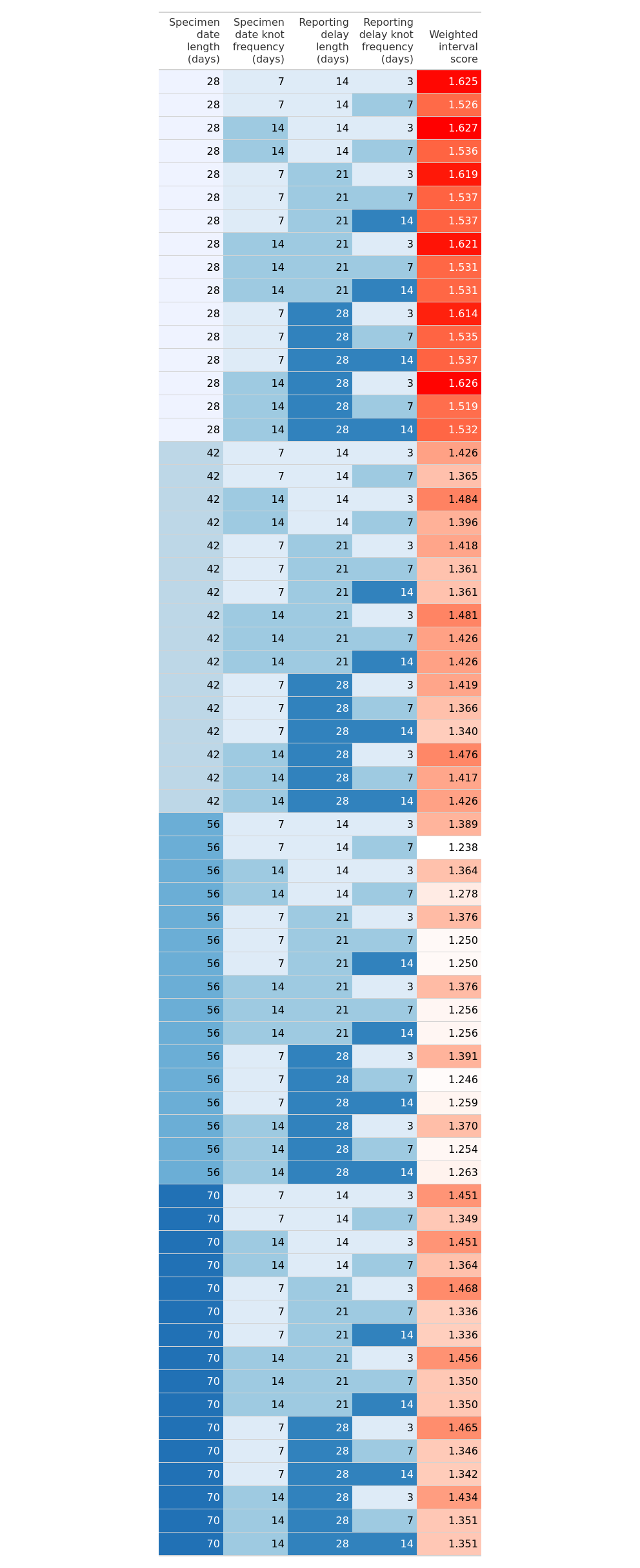

Supplementary Table 2. Average daily scores over the tuning period for the GAM model for different values of hyperparameters.

**Section 3 - epinowcast**

We tuned two parameters: the maximum number of days to model in the delay distribution and the training length (number of days of data used to nowcast). Based on descriptive analysis of empirical reporting delays we tested maximum delays between 14 and 28 days, as well as training lengths between 21 and 49 days, shown in Supplementary Table 3. We found a maximum delay of 7 and a training length of 35 was optimum. Priors were specified to be informative, using the descriptive analysis available from the data, outlined in Supplementary Table 4. We did not tune priors to improve scoring, but rather selected sensible parameters based on known information before observing scoring results. A day of week effect for the hazard function was explored, but did not converge for all tuning results, nor did a day of week effect in the reference model converge.

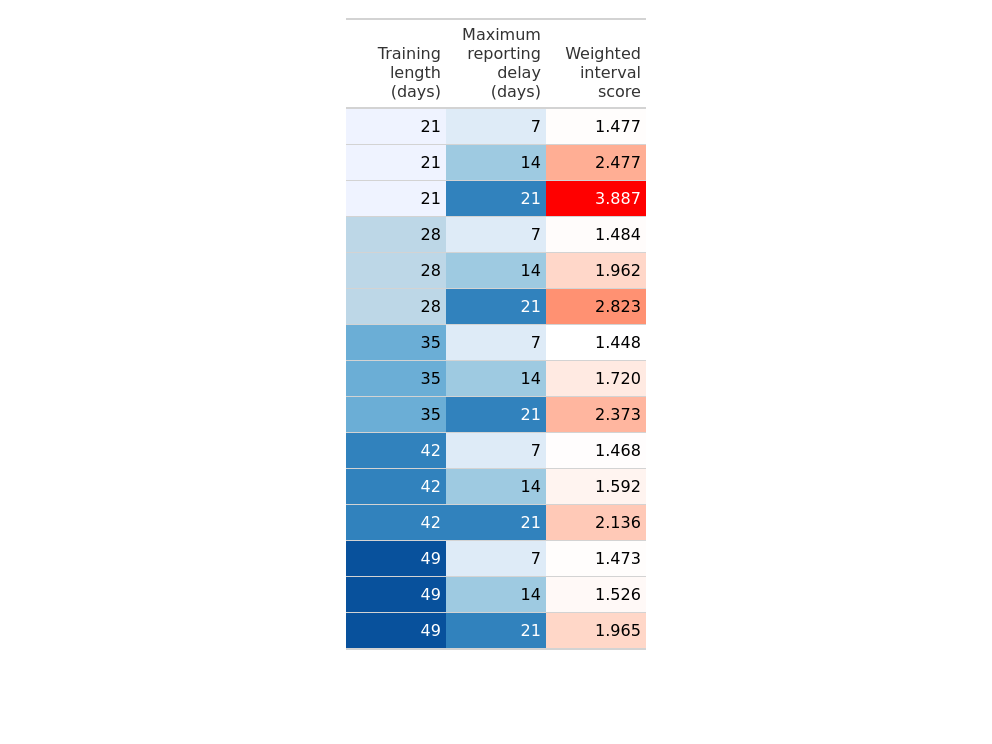

Supplementary Table 3. Average daily scores over the tuning period for the epinowcast model for different values of hyperparameters.

| **Parameter Description** | **Distribution** | **Mean** | **Standard deviation** |
| --- | --- | --- | --- |
| Log mean intercept for parametric reference date delay | Normal | log(3) | 0.2 |
| Log standard deviation for the parametric reference date delay | Zero truncated normal | 0.3 *  sqrt(log(1)-log(5)) | 0.2 |
| Standard deviation of scaled pooled parametric mean effects | Zero truncated Normal | 0.0 | 0.2 |
| Standard deviation of scaled pooled parametric standard deviation effects | Zero truncated Normal | 0.0 | 0.2 |

Supplementary Table 4. Priors specified for epinowcast model.

**Section 4**

**BSTS**

We use inverse Gamma distributions for variance as priors for the standard deviations $\sigma_{\mu}$ and $\sigma_{\delta}$, specifying an upper limit for in the *sd.prior* function. The standard deviation for seasonality $\sigma_{\tau}$ is selected by the *bsts* function. Posterior samples are generated for 7 days into the future, with prediction intervals created using quantiles on the posterior samples. Hyperparameters are chosen to optimise model performance on the forecasted 7 days, based on averaged daily scores (Supplementary Table 3). We choose a training length of 60 days so that the model adapts to recent data, and an upper limit for $\exp(\sigma_{\mu})$ of 1.1 and $\exp\left( \sigma_{\delta} \right)$ of 1.1; we anticipate that the subsequent period will have greater variability compared to the tuning period and these values correspond to a 10% change in the mean and a daily accumulating 10% change in the slope respectively.

**BSTS + NHS 111 online**

For the “BSTS + NHS 111 online” model we choose a training length of 150 days, an expected model size of 5 and an upper limit for $\exp(\sigma_{\mu})$ of 1.01 and $\exp\left( \sigma_{\delta} \right)$ of 1.1, which correspond to a 1% change in the mean and a daily accumulating 10% change in the slope respectively. Scoring across each parameter are shown in Supplementary Table 4.

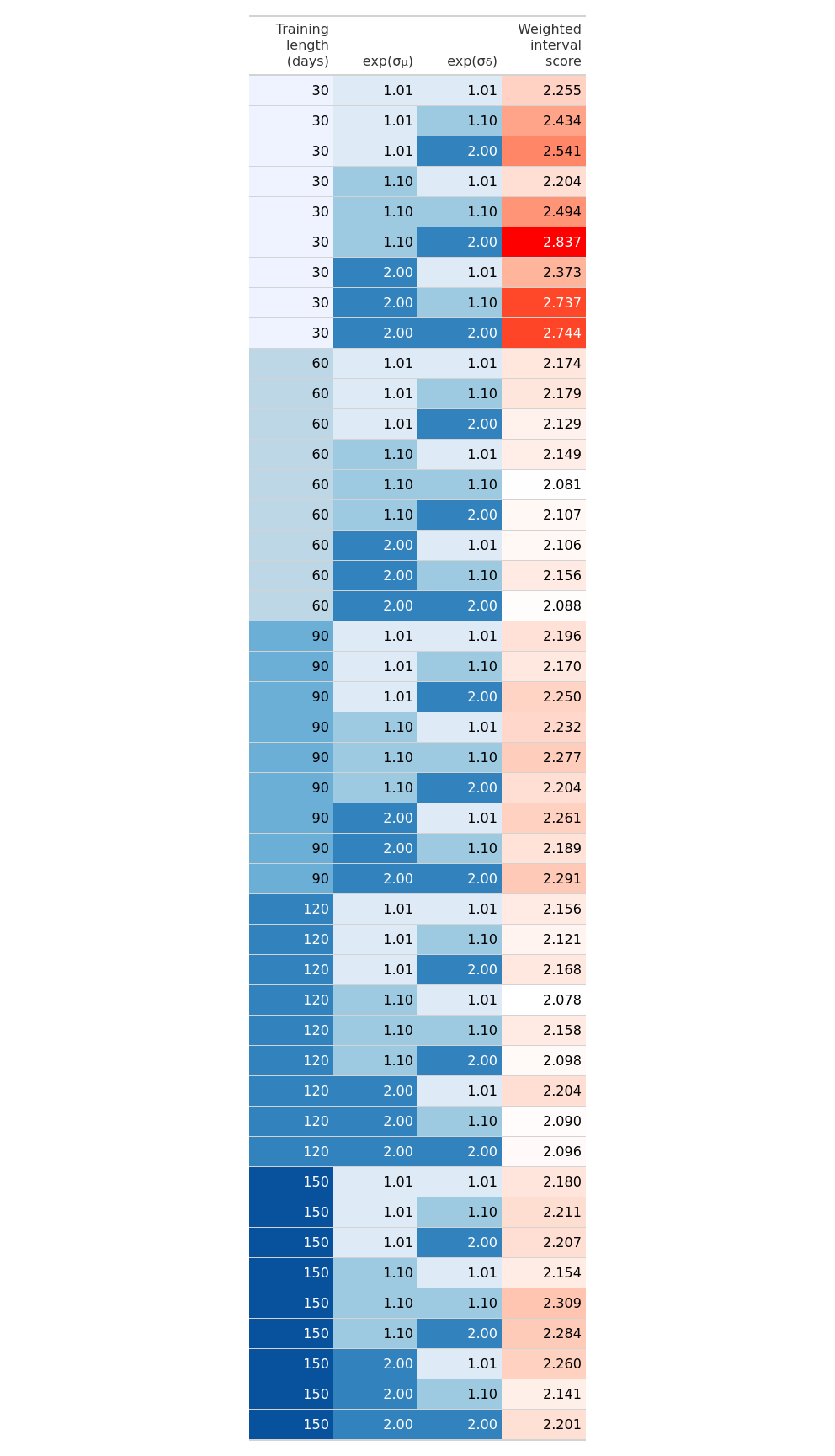

*Supplementary Table 5. Average daily scores over the tuning period for the BSTS model by training length and* $\sigma_{\delta}$ *and* $\sigma_{\mu}$ *values.*

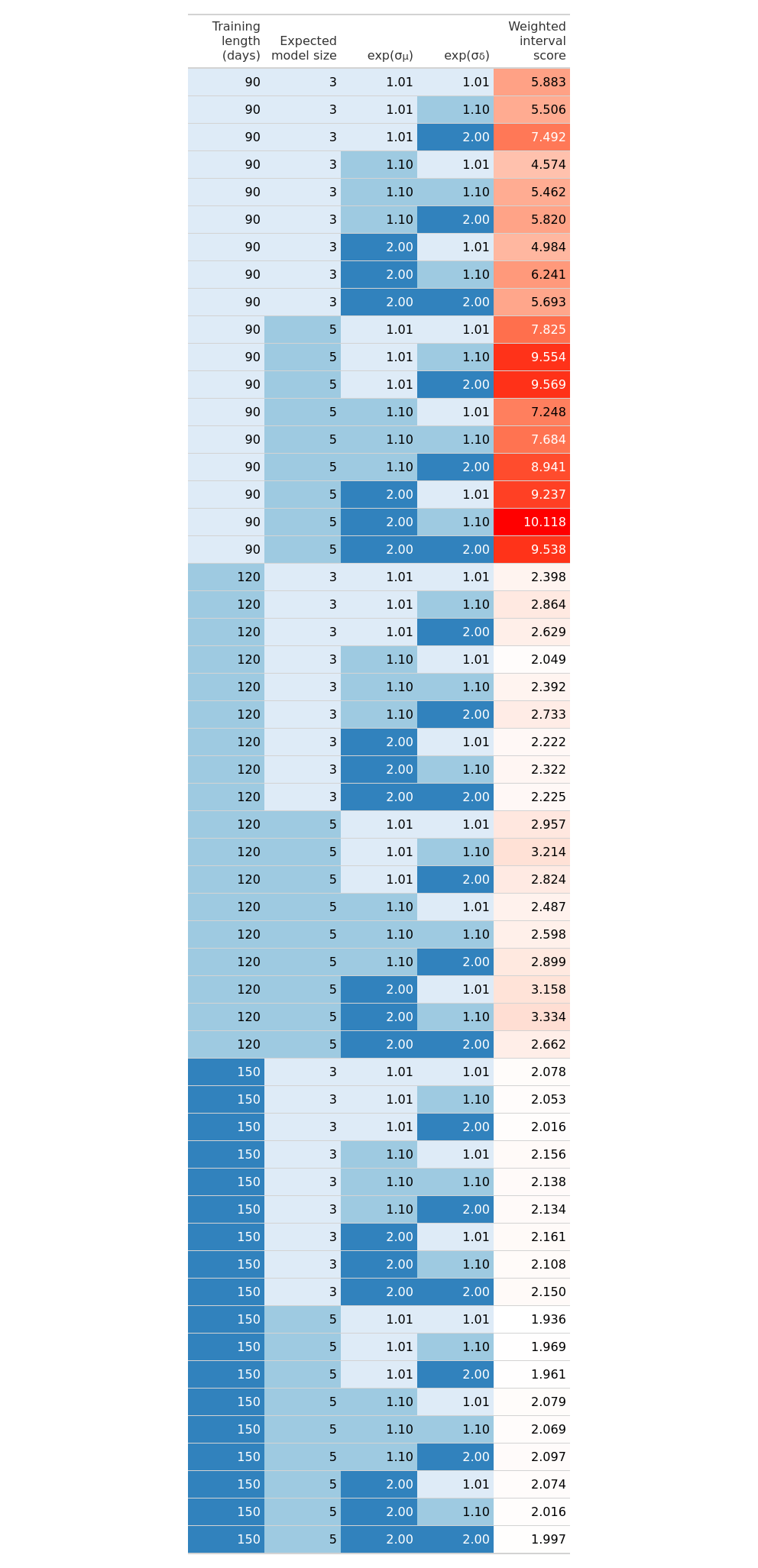

*Supplementary Table 6. Average daily scores over the tuning period for the BSTS + NHS 111 online model by training length, expected model size and* $\sigma_{\delta}$ *and* $\sigma_{\mu}$ *values.*

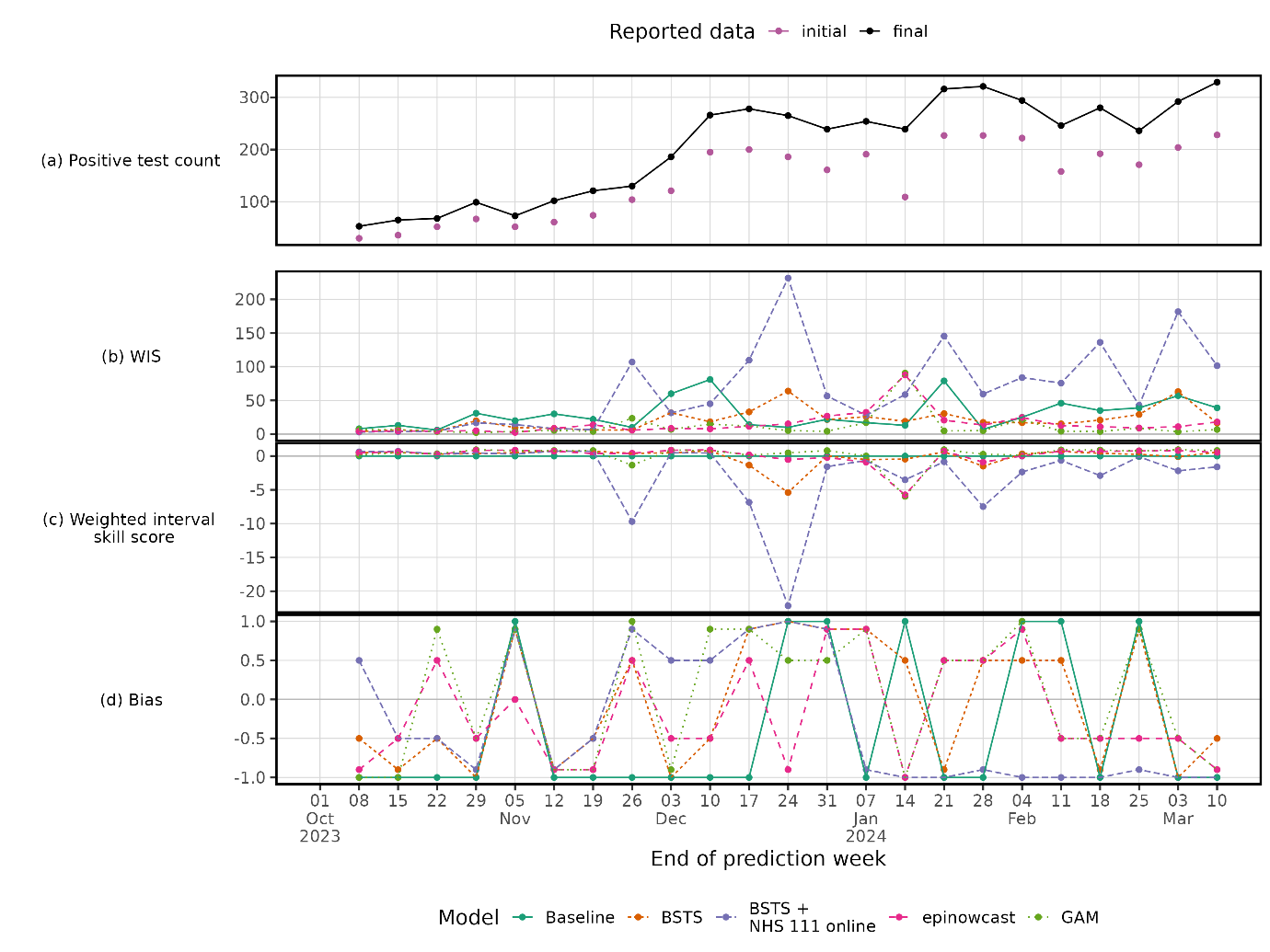

Supplementary Figure 5. Weekly count of final and initial reported tests (top pane) with weekly model scores.

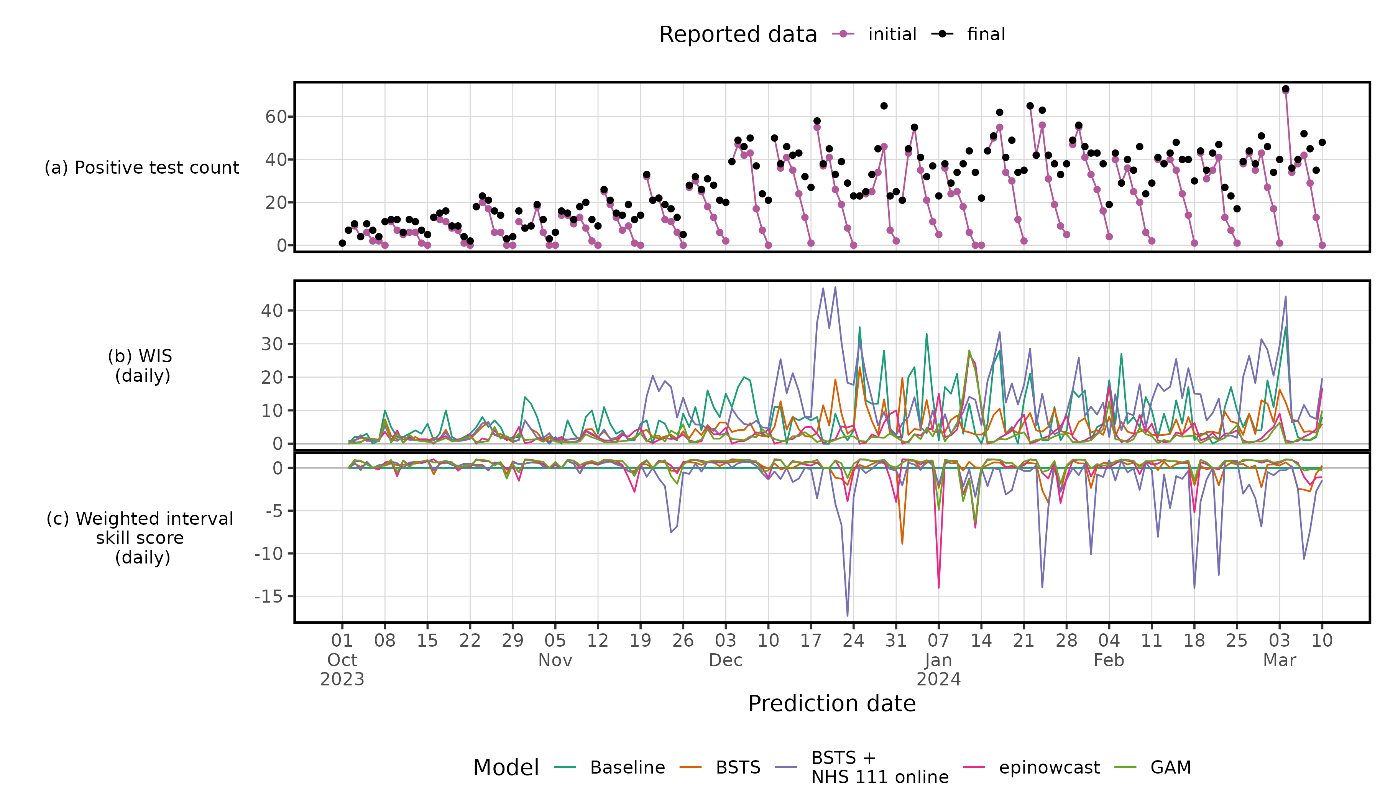
 Supplementary Figure 6. Daily count of final and initial reported tests (top pane) with daily model scores.
